# Paediatric Attendances and Acuity in the Emergency Department during the COVID-19 Pandemic

**DOI:** 10.1101/2020.08.05.20168666

**Authors:** Katy Rose, Kerry Van Zyl, Rachel Cotton, Susan Wallace, Francesca Cleugh

**Affiliations:** Paediatric Emergency Department, Imperial College Healthcare NHS Trust, London, UK; Paediatric Department, Chelsea and Westminster NHS Foundation Trust, London, UK

**Keywords:** Paediatrics, Emergency Department, Patient Acuity, Health Service Needs and Demands, Pandemics

## Abstract

**Aim:** To investigate the difference in both numbers and acuity of presentations to the Paediatric Emergency Department (PED) during the peak time period of the current global SARS-CoV-2 pandemic.

**Design:** This single centre retrospective observational study used routinely collected electronic health data to compare patient presentation characteristics between 21^st^ March and 26^th^ April 2020 compared to the equivalent time period in 2019.

**Results:** There was a 90% decrease in attendances to PED, with a 10.23% reduction re-attendance rate. Children presenting were younger during the pandemic, with a median age difference of 2 years. They were more likely to present in an ambulance (9.63%), be admitted to hospital (5.75%) and be assigned the highest two Manchester triage categories (6.26%). There was a non-significant trend towards longer lengths of stay. The top 10 presenting complaints remained constant (although the order changed) between time periods. There was no difference in mortality or admission to PICU.

**Implications:** Our data demonstrates that there has been a significant decrease in numbers of children seeking emergency department care. It suggests that presenting patients were proportionally sicker during the pandemic; however, we would argue that this is more in keeping with appropriate acuity for PED presentations, as there were no differences in PICU admission rate or mortality. We explore some of the possible reasons behind the decrease in presentations and the implications for service planning ahead of the winter months.

## INTRODUCTION

On 11^th^ March 2020 the WHO declared the COVID-19 outbreak (the disease caused by SARS-CoV-2) a pandemic^1,2^. In keeping with global epidemiology, in the UK, the disease has predominantly affected older patients, with only one death below 14 years of age^3^. Due to rapidly evolving testing criteria, the peak number of infections remains unknown. The highest daily COVID-19 attributable death toll in England and Wales occurred on 8^th^ April 2020^3^.

There has been a 22% increase in attendance rates to the Emergency Department (ED) across the UK since 2008/9^4^. Proportionally, paediatric patients attend ED more frequently than adults^5^. In 2016/17, there were 425 ED attendances for every 1,000 children/young people compared with 345 ED attendances for every 1,000 adults aged over 25^6^.

Since the pandemic was declared there has been a sharp fall in UK ED attendances compared to previous years (−30% in March and −56% in April)^4,7^. This trend has been noted globally, but it remains unclear why ^8–11^. Age divided data is not collected nationally but there is growing concern that the fall in paediatric attendance is even starker and possibly as high as 90%^8,12^.

This study compares the routinely collected attendance data of children presenting to a central London teaching hospital Paediatric Emergency Department (PED) during the peak weeks of the pandemic in the UK to an equivalent period in 2019. We explore differences in baseline presentation data, admission rates and length of stay and discuss possible underlying reasons. We also examine the impact of changes to PED services as a result of the pandemic.

## METHODS

### Study Design and Inclusion Criteria

This retrospective observational cohort study from a single central London PED compared attendances between 21^st^ March 2020 and 26^th^ April 2020 and of the equivalent period in 2019. The department routinely sees all patients up to 16 years old; although some young people with chronic conditions are seen until they transition to adult services, they were excluded from this analysis.

During the pandemic, the department was divided using temporary walls into COVID-19 “red” high risk) and “green” (low risk) areas. Patients presenting with any of fever >37.5°C, cough, shortness of breath, flu-like symptoms or diarrhoea were streamed into the “red” area; all others were assessed in “green”. All patients being admitted were tested for SARS-CoV-2.

### Data Collection

The Qlikview analytics platform was used to interrogate electronic health records. Extracted data included baseline patient demographics (age and gender) and attendance characteristics (Manchester triage category^13^, mode of presentation, referral source, presenting complaint category and discharge destination). Admission numbers and length of stay as collected by paediatric wards and the clinical decisions unit (CDU) was also extracted.

Ward-collected length of stay data for 2019 included patients admitted for ambulatory review or appointments (such as phlebotomy or for single antibiotic doses). These services were largely moved to alternative locations during the pandemic. To prevent unrealistic skew, admissions shorter than 4 hours were excluded from calculation of median length of stay for both time periods. CDU length of stay data was excluded as the unit was closed during the pandemic.

This study was reviewed and approved by the local Audit and Service Evaluation Board prior to commencement (Imperial College Healthcare, UK; registration number: 487).

### Statistical Analysis

Univariate analysis and data visualisation to include frequency, percentages, medians and means were performed in Microsoft Excel^14^. R Studio was used to evaluate significance of difference in proportions using the two-tailed Z-test, calculate binomial confidence intervals using the Clopper-Pearson exact method, and compare differences in non-parametric data (age and length of stay) using the Mann-Whitney U test^15^.

## RESULTS

There were 453 patient attendances (376, 83.2% unique attenders) between 21^st^ March and 26^th^ April 2020; the same period in 2019 saw 4238 attendances (3092, 73.0% unique attenders), representing an 89.3% reduction (table 1). The decrease in rate of unique attendances was significant, with higher rates during the COVID-19 period (10.2%, Cl 6.28 – 13.7%, p<0.001). During the pandemic period only 5 (1.10%) patients re-attended more than once compared to 89 (2.10%) during the 2019 time period (p=0.21).

**Table 1.**
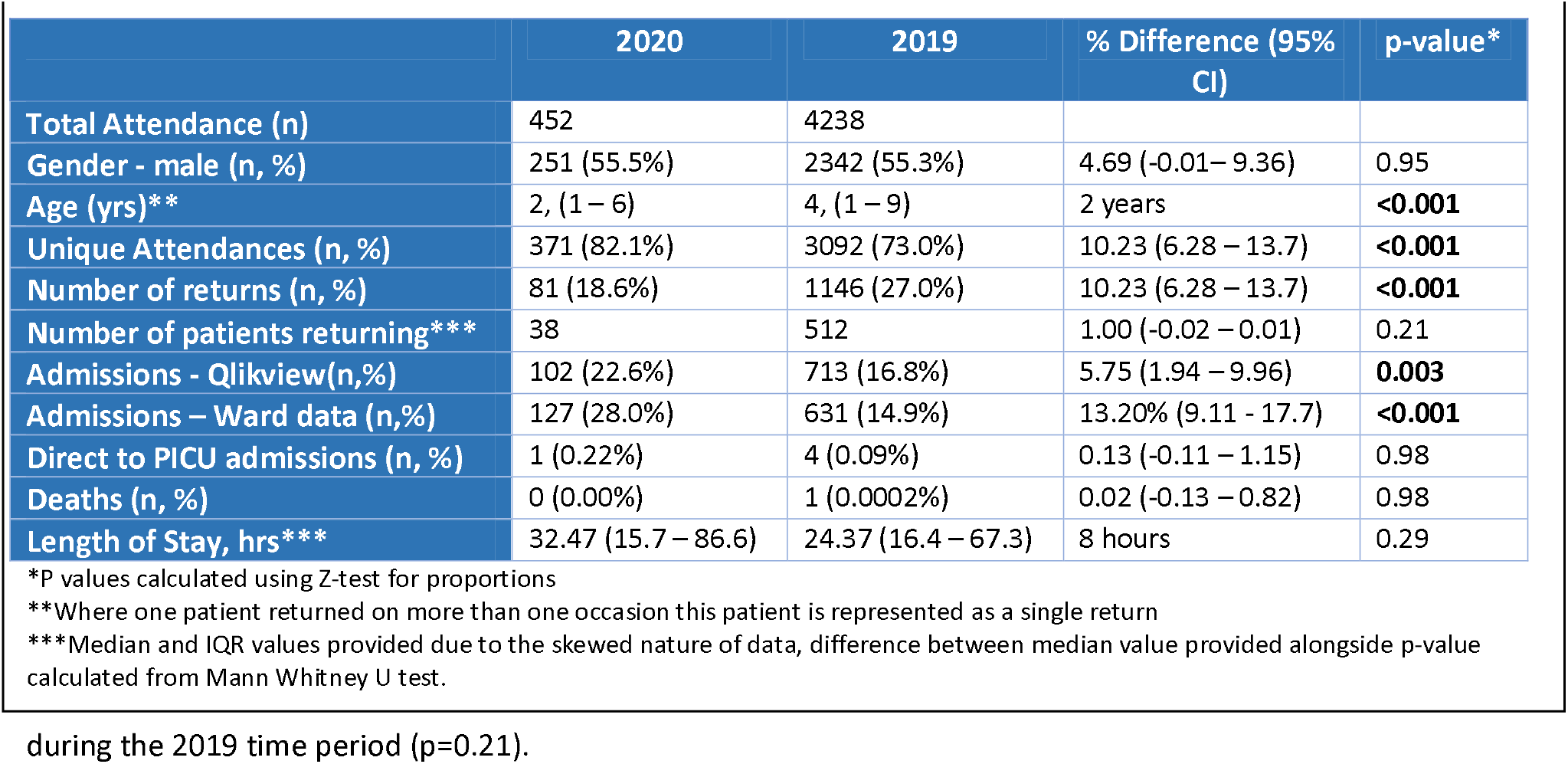
Demographics and admission characteristics.

During the pandemic period, 46.2% of patients were assessed in the “red” area, and 43.8% in the “green”. The remaining 9.95% were directed to other services outside the emergency department.

There was a younger median patient age (2yrs, IQR 1-6) during the pandemic compared to the previous year (4yrs, IQR 1-9), (p<0.001). Attendance rates by gender were similar between the two time periods (p=0.95).

Admission rates direct to PICU were very low during both periods. During the pandemic, 102 patients were coded as admitted from the Qlikview platform, compared to 713 patients during the 2019 period, representing an increased admission rate of 5.75% (Cl 1.94 – 9.96%, p=0.003). Ward-collected data demonstrated 127 (28.0%) admissions during the pandemic and compared to 631 (14.9%) admitted to the ward/CDU during 2019, representing an increased admission rate of 13.2% (Cl 9.11-17.7%, p<0.001).

Median length of ward stay increased by 8 hours during the pandemic to 32.5 hours (IQR 15.6 – 67.0) from 24.37 (IQR 16.4 – 67.3) (p=0.29).

5 admitted patients had a positive SARS-CoV-2 test (4.90% of total admissions). Of these, two were neonates (under 28 days), two were under 2 months old, and one was 7-years old. Four of the SARS-CoV-2 positive patients were male, four were admitted to the General Paediatric ward and only one patient required PICU level care. One patient had respiratory symptoms at presentation and four patients had fever. Only one patient had a primary diagnosis of COVID-19, and they did not require PICU care.

The top 10 presenting complaint categories remained the same during pandemic and 2019 periods (table 2, figure 1), accounting for 73.7% and 72.3% of attendances respectively (p=0.57). However, within these top 10 categories, the order was different. In particular, there was a higher proportion of unwell new-borns (+4.98%, Cl 2.71 – 7.91%, p<0.001) and unwell babies (+6.29%, Cl 3.72 – 9.50%, p<0.001) and a lower percentage of limb problems (−7.50%, Cl −9.24 – −5.16%, p<0.001).

**Figure 1:**
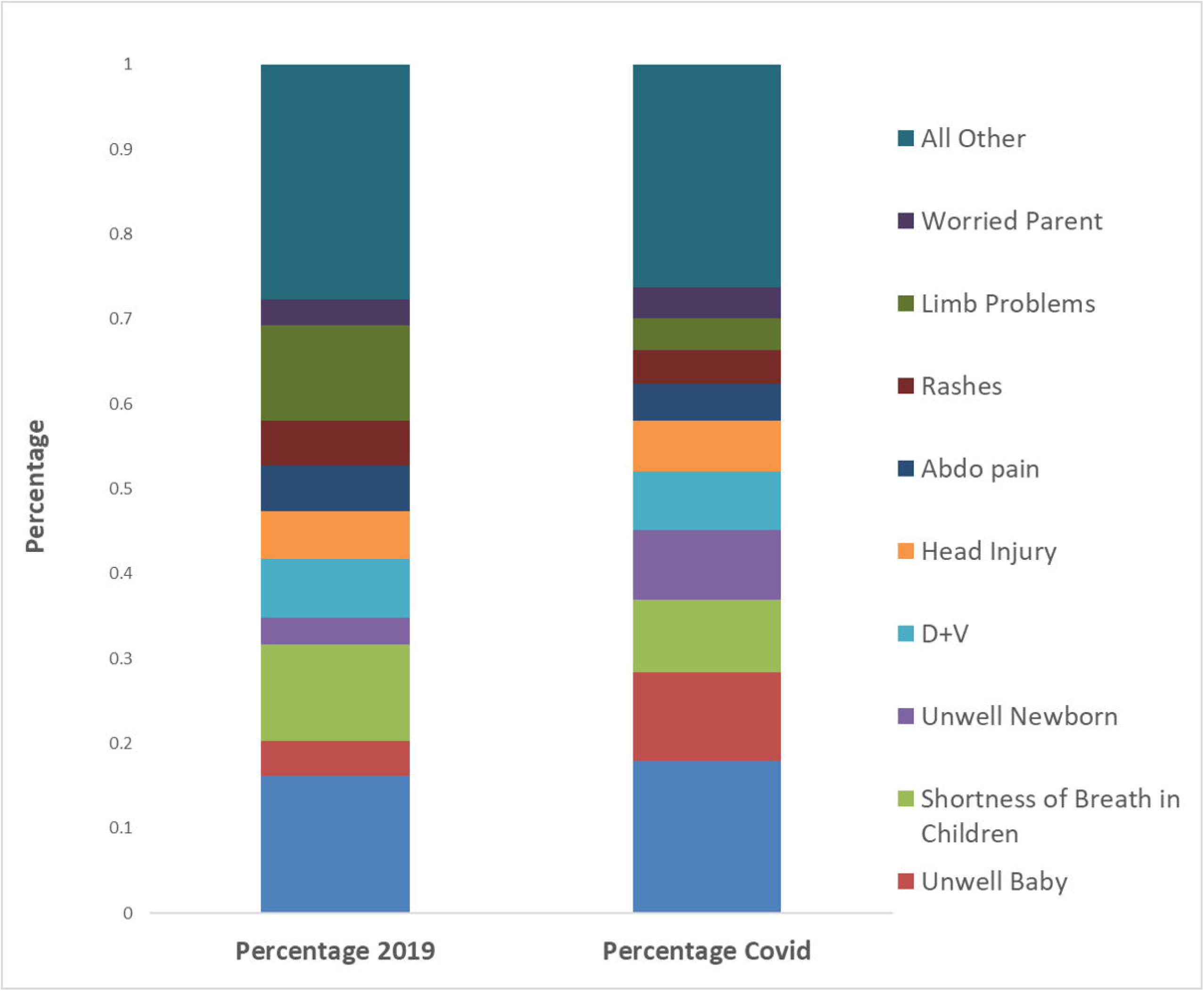
Top 10 Presenting Complaints.

**Table 2:** Top 10 Presenting Category.

| **Table 2: Top 10 Presenting Category** |  |  |  |  |
| --- | --- | --- | --- | --- |
|  | **2020 N (%)** | **2019 N (%)** | **% Difference (95% CI)** | **p-value\*** |
| Unwell Child | 81 (17.9%) | 687 (16.2%) | 1.71 (-1.75 – 5.66) | 0.37 |
| Unwell Baby | 47 (10.4%) | 174 (4.11%) | 6.29 (3.72 – 9.50) | **<0.001** |
| Shortness of Breath in Children | 39 (8.63%) | 479 (11.3%) | -2.67 (-5.15 – 0.41) | 0.1 |
| Unwell New-born | 37 (8.19%) | 136 (3.21%) | 4.98 (2.71 – 7.91) | **<0.001** |
| D+V | 31 (6.86%) | 291 (6.87%) | -0.01 (-2.16 – 2.79) | 1 |
| Head Injury | 27 (5.97%) | 240 (5.66%) | 0.31 (-1.68 – 2.96) | 0.87 |
| Abdominal pain | 20 (4.42%) | 227 (5.36%) | -0.93 (-2.65 – 1.46) | 0.46 |
| Rashes | 18 (3.98%) | 225 (5.31%) | 1.33 (-2.98 – 0.98) | 0.27 |
| Limb Problems | 17 (3.76%) | 477 (11.3%) | -7.50 (-9.24 – -5.16) | **<0.001** |
| Worried Parent | 16 (3.54%) | 128 (3.02%) | 0.52 (-0.95 – 2.70) | 0.64 |
| All Other | 119 (26.3%) | 1174 (27.7%) | -1.37 (-5.47 – 3.07) | 0.57 |

\*P values calculated using Z-test for proportions

During the pandemic, there was no significant difference in patients triaged to the highest Manchester triage category (p = 0.97). There was a significant increase in patients assigned the second highest Manchester triage category (+6.12%, Cl 2.55 to 10.2%, p < 0.001). Concordantly, there was a significant decrease in category green patients (−5.76%, Cl −10.6 to – −0.92%, p = 0.02). Although there was a significant increase in category blue patients (+1.11%, Cl 0.19 – 2.80%, p = 0.022), absolute category blue patient numbers were low in both time periods (pandemic: 9, 2019: 28) (table 3, figure 2).

**Figure 2:**
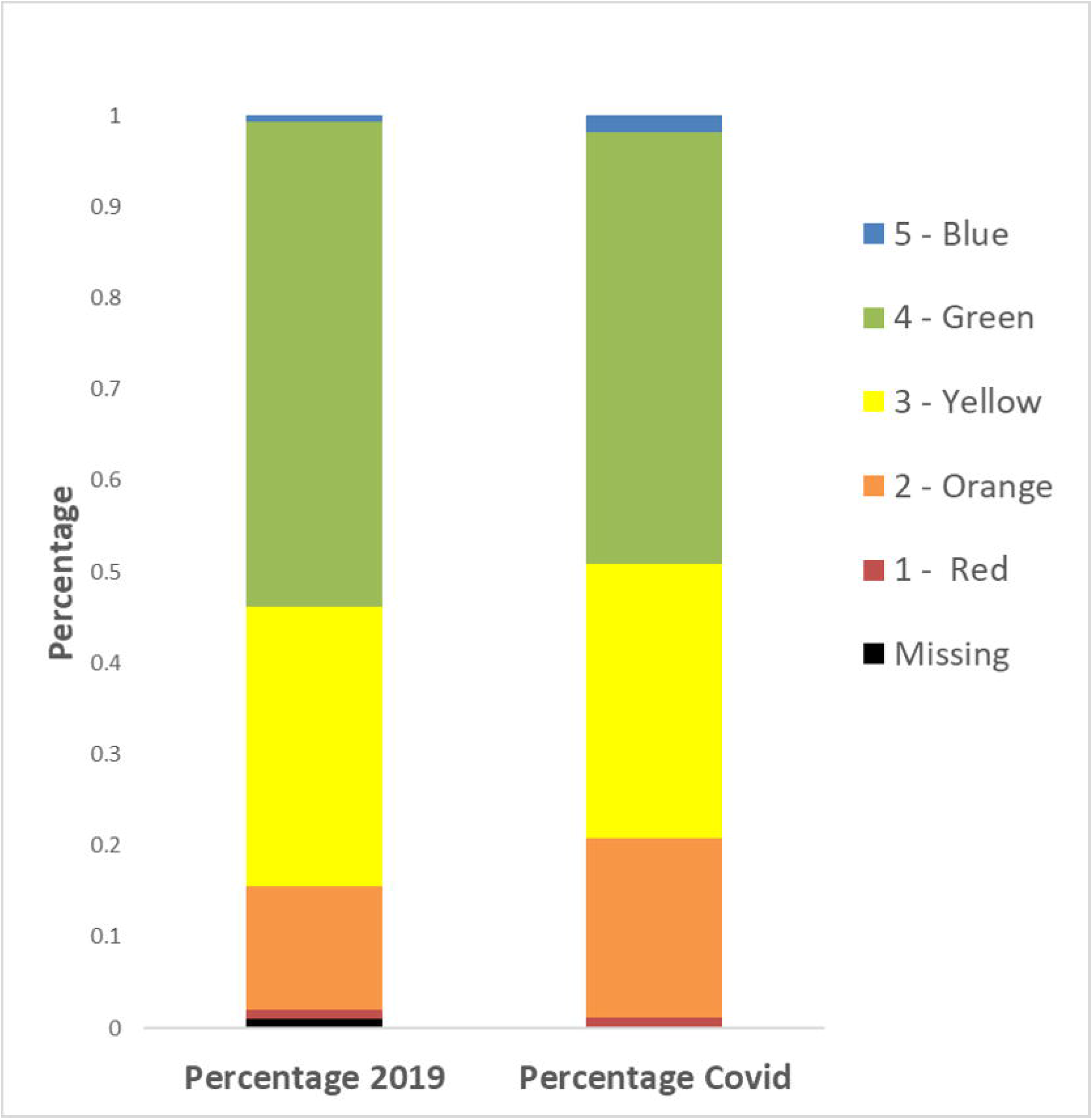
Triage Category at Arrival.

**Table 3:** Triage Categories at Arrival.

|  | 2020 N (%) | 2019 N (%) | % Difference (95% CI) | P-Value* |
| --- | --- | --- | --- | --- |
| Missing | 0 (0.00%) | 42 (0.99%) | -0.99 (-1.34 – -0.15) | 0.062 |
| 1 - Red | 5 (1.11%) | 41 (0.97%) | 0.14 (-0.59 – 1.61) | 0.97 |
| 2 - Orange | 89 (19.7%) | 575 (13.6%) | 6.12 (2.55 – 10.22) | **<0.001** |
| 3 - Yellow | 136 (30.1%) | 1301 (30.7%) | -0.61 (-4.90 – 3.98) | 0.83 |
| 4 - Green | 214 (47.4%) | 2251 (53.1%) | -5.76(-10.6 – -0.92) | **0.022** |
| 5 - Blue | 8 (1.77%) | 28 (0.66%) | 1.11(0.19 - 2.80) | **0.022** |
| Top two Triage Categories | 94 (20.8%) | 616 (14.5%) | 6.26 (2.60 – 10.4) | **<0.001** |
| Bottom two Triage Categories | 222 (49.1%) | 2279 (53.8%) | -4.66 (-9.48 – 0.18) | 0.066 |

\*P values calculated using Z-test for proportions

During the pandemic children were significantly more likely to present in an ambulance (+9.63%, Cl 6.38 – 13.4%, p<0.001), be referred back to ED from the Urgent Care Centre (UCC) (+6.27%, Cl 3.96 – 9.25, p<0.001), or be referred from another healthcare service (+1.56%, Cl 0.22 – 3.60%, p<0.001) (table 4). In 2019, patients were significantly more likely to be referred by their GP (−2.56%, Cl −3.61 to −0.87%, p=0.01) or parent/self-present (−15.1%, Cl −19.7 to −10.8%, p<0.001). There was no significant difference in referrals from the 111 service, Urgent Care Services or non-health institutions (table 4).

**Table 4:** Referral Sources.

|  | 2020 N (%) | 2019 N (%) | % Difference (95% CI) | P-Value* |
| --- | --- | --- | --- | --- |
| GP | 7 (1.55%) | 174 (4.11%) | -2.56 (-3.61 – -0.87) | **0.01** |
| Parents/self | 307 (67.9%) | 3518 (83.0%) | -15.09 (-19.7 – -10.8) | **<0.001** |
| 111 | 1 (0.22%) | 13 (0.31%) | -0.086(-0.38 – -0.94) | 1 |
| UCC | 4 (0.88%) | 61 (1.44%) | -0.56(-1.25 – 0.84) | 0.46 |
| Emergency Services | 75 (16.6%) | 295 (6.96%) | 9.63 (6.38 – 13.4) | **<0.001** |
| Other healthcare service provider | 14 (3.10%) | 66 (1.56%) | 1.54 (0.22 – 3.60) | **0.027** |
| A - UCC Return | 39 (8.63%) | 100 (2.36%) | 6.27 (3.96 – 9.25) | **<0.001** |
| Other non-health institution | 5 (1.11%) | 11 (0.26%) | 0.85 (0.18 – 2.31) | 0.52 |

\*P values calculated using Z-test for proportions

## Discussion

We found that, during the pandemic, number of presentations to PED has significantly reduced. A significant professional concern was whether the reduction in attendance may represent a failure of patients to seek appropriate healthcare, leading to late presentations of more unwell children^12^. Emerging evidence in the UK is more reassuring^16^. In our study, those presenting tend to be sicker (as per mode of presentation, Manchester triage category and ward admission rates), although there was no significant difference in median length of ward stay or PICU admission rate.

The use of routinely collected data allowed us to explore whether the fears of a late-presenting and sicker cohort of children had basis, both to help inform preparation for a second wave, and try to explore changes in health seeking behaviour. At pandemic onset acute pathways altered to streamline services and minimise exposure risk. Several services operated ‘direct to specialist’ review which contributed to decreased PED footfall. A separate, nearby ‘green’ hospital was created to accept direct primary care referrals and to replace PED review clinics; this likely reduced footfall through PED. Conversely, this involved closing a walk-in service, possibly increasing study PED attendances.

As expected from the epidemiology of the SARS-CoV-2 virus, few children were admitted with a positive SARS-CoV-2 PCR test; background carriage rate remains unknown due to testing policies. SARS-CoV-2 has not severely affected children, however there have been decreases in presentations to paediatric healthcare services^8,10^’^12^’^17^. The observed decrease in our study was nearly 90%. Of those who attended roughly 45% were “red” requiring isolation. This became achievable because of the low overall numbers despite minimal isolation spaces. Maintenance of social distancing was also possible because of low attendance numbers.

Preparing for winter months, maintaining these changes to help ease pressures on PEDs will be important. Few, if any, PEDs have capacity to isolate all ‘red’ patients and ensure social distancing with normal attendance figures.

The study centre is a level 1 trauma centre in a major metropolitan area; the reduced local footfall caused by ‘lock-down’ may explain the decrease. Limitation on the spread of other infectious agents, coupled with a reduction in risk inherent activities (e.g. team sports and road use) will also have contributed. Parental use of 111 and online information may also have reduced unnecessary attendance. The need to understand these public health factors will become crucial to managing the busier winter period ahead; potentially preventable hospital attendances may be far greater than previously appreciated^16^.

Our data supports the concern that, during the pandemic, children who present are sicker, but they are not in extremis. There were more children conveyed by ambulance and a higher proportion of PED attenders who were assigned higher overall triage categories. However, there was no difference in proportion at the highest “Red” triage category – defined as those who need care immediately. Reassuringly, death rates in the PED remained very low; no deaths occurred during the pandemic period. Additionally, direct admission to PICU was no different from 2019. We argue that changes in other acuity markers actually represent a more appropriate utilization of PED.

Of interest was the proportion of unwell new-borns, a vulnerable population, presenting during the pandemic, 10.4%, when compared to 2019, 3.21%. Further work would be required to quantify the direct impact of reduced community services in the UK in response to the pandemic, which has been implicated to factor internationally for increased neonatal presentations ^18^.

We noted proportionally more ward admissions during the pandemic. The decision to admit patients to hospital is multifactorial,^19–21^ with a wide range of admission appropriateness amongst paediatric patients^22–26^. In context of appropriate admissions, the UK performs particularly well in the global setting^22^. It is also suggested that in busier departments rates of admission increases ^27,28^. Given the significant fall in patient numbers, our suggestion is that the reflected higher ward admission rate is more likely to represent a slightly sicker patient cohort compared to 2019. It is difficult to determine how factors such as decreased bed occupancy and increased resource availability at initial assessment, including diagnostic imaging or neurophysiology, may have influenced rates.

This study has some limitations. There are known strengths and limitations to using routinely collected information from electronic health records (EHR)^29–31^. The Qlikview platform derives admission rates based on outcome decisions coded in PED. The paediatric wards and CDU both keep independent data sets of admissions. There were discrepancies between the two. The Qlikview platform admission rate may appear elevated because coding occurs at the point a bed is booked; it is not uncommon for alternative pathways to become available in light of late test results, specialist reviews and availability of community services. Ward-collected admissions may have appeared elevated because PED is not the only pathway for paediatric admissions. In including both calculations, we aimed to overcome this limitation.

The choice of studied outcomes was restricted by the maintained databases. They included standard proxy markers for acuity including Manchester triage score, general and intensive care admissions, mortality and length of hospital stay. We also included the mode of presentation; although there are a proportion of likely inappropriate use of ambulances this is lower than the general inappropriate attendance rate to ED^32–35^. Although studied outcomes are limited, we feel that they are sufficient to start understanding the changes observed during the pandemic. We hope that these insights can act as springboards for further work both locally and nationally.

Increasing numbers of hospitals have access to EHR. We would encourage other groups to look at how their population’s behaviour altered. One of the key benefits to this study design is the speed; our overarching aim must be that these efforts can be co-ordinated nationally to help contribute to public health planning. This includes short-term planning ahead of the coming winter, but also thinking longer term about how acute paediatric care is delivered, and whether more PED visits are preventable.

Judging the appropriateness of presentations to both adult and paediatric ED remains contentious ^33,36–38^. This was not the focus of this evaluation but should be highlighted given the magnitude of changes in attendance rates. Continued evaluation of service use and exploration of the lessons of the early pandemic may help to reduce the burden on EDs.

## Conclusion

This study demonstrated a significant decrease in paediatric attendances to the PED at the peak of the COVID-19 pandemic. Although presenting patients may have been sicker than pre-pandemic levels, there is no evidence that this had negative effects on immediate patient outcomes. It is unclear why this effect was seen. Further research around changes to disease, community services and injury profiles caused by isolation and activity restrictions is required. Additionally, work with parents and children to understand decisions around healthcare utilization during the pandemic is vital.

If a second wave of COVID-19 occurs it seems likely that policies developed for the initial pandemic remain appropriate; concern about acute paediatric outcomes can, in part, be allayed. A key remaining concern is whether departments will be able to maintain appropriate isolation policies in the usually busier winter months, especially if social isolation and activity levels are not decreased as during the peak of the pandemic. Engaging in early planning will be essential.

Funding statement: This research received no specific grant from any funding agency in the public, commercial or not-for-profit sectors.

Contributions: K.R and S.W conceived and planned the study with supervision and support from F.C. K.R, R.C and K.VZ carried out data collection. K.R and K.VZ contributed to the interpretation of the results. K.R. took the lead in writing the manuscript. All authors provided critical feedback and helped shape the research, analysis and manuscript.

What is already known:

- The SARS-CoV-2 virus causes a much more prevalent serious illness in adults compared to children
- Attendance rates to emergency departments has decreased across locations and presentation type
- Large scale service changes occurred across the NHS to help prepare for predicted demand for adult critical care provision

What this paper adds:

- Attendance rates to this paediatric emergency department was much lower than the overall data across ages
- Children may appear slightly sicker at presentation but there was no associated increase in mortality or PICU requirement in the ED
- Without this significant decrease in attendance rates PEDs face challenges to attain necessary isolation and social distancing requirements without substantial changes in service delivery

## Data Availability

Original data can be made available at request

